# Cytotoxic and regulatory CD8 T cells dynamics underlies ICI-myotoxicity outcome

**DOI:** 10.64898/2026.03.03.26347221

**Authors:** Runci Wang, Chunyan Xiang, Adrien Procureur, Julian Sanchez-Dal Cin, Soon-min Hong, Boyang Zhang, Xiaolin Lin, Xinyue Lian, Guangcheng Liu, Wu Wei, Xueqin Chen, Xiuying Xiao, Xiaoxiang Chen, Xiaodong Wang, Michelle Rosenzwajg, Yves Allenbach, Qiong Fu, Nan Shen, Joe-Elie Salem, Shuang Ye

## Abstract

Immune checkpoint inhibitor–induced myotoxicity (ICI-M) comprises myocarditis, myositis, and myasthenia gravis–like syndrome, demanding rapid recognition and therapy. Using immunophenotyping and transcriptomics analysis from blood and muscle, we identified distinct CD38^hi^ and KIR^+^ CD8 T cells in ICI-M. Abatacept rescued patients and altered the composition and clonality of these cells. Dynamics of CD38^hi^ and KIR^+^ CD8 T cells effectively supported therapeutic monitoring, offering personalized treatment in life-threatening irAEs.

## Introduction

Immune checkpoint inhibitors (ICI) block CTLA-4 or PD-1/PD-L1 to enhance CD8 T cell-mediated anti-tumor immunity^1,2^ but frequently trigger immune-related adverse events (irAEs) in over 80% of patients^3^. ICI myotoxicity (ICI-M) present days to weeks after ICI initiation as a life-threatening triad of myositis, myocarditis, and myasthenia gravis-like syndrome^4^, requiring prompt recognition and tailored immunosuppression to prevent mortality.

Effective management of fulminant ICI-M remains an unmet clinical need. Abatacept, a CTLA-4 agonist that inhibits CD80/86-CD28 co-stimulation, combined with the JAK1/2 inhibitor ruxolitinib, has shown promise in treating glucocorticoid-refractory or fulminant ICI-M, suggesting a previously underappreciated decelerating effect on immune activation^5–7^. Real-time monitoring of cellular responses to such therapy could guide treatment decisions and reveal underlying mechanisms.

Shared CD8 T cell clonotypes have been found in muscle, myocardium, and tumor tissue of ICI-M patients, indicating a delicate dysregulation between anti-tumor immunity and autoimmunity^8^. T cell clones were reported to target alpha-myosin in mice model of ICI-M while most autoantigens remain unknown^9,10^. Cytotoxic CD8 T cells were also clonally expanded in the peripheral blood of patients with ICI-myocarditis^11^. Previously we identified a clonal CD38^hi^ CD8 T cell population directly bound and activated by anti-PD-1 antibodies and specifically expanded in the blood and joints in ICI-arthritis (ICI-A), indicating an irAE specific T cell activation in ICI-toxicity^12^. Additionally, regulatory CD8 T cells expressing killer cell immunoglobulin-like receptors (KIRs) modulate autoreactive T cells in autoimmunity and cancer^13,14^. Whether these populations facilitate clinical recognition and therapeutic monitoring in ICI-M requires investigation.

To dissect the immunological mechanism underlying ICI myotoxicity and its treatment, we used multiparametric flow cytometry, single-cell RNA sequencing (scRNA-seq) and T cell receptor (TCR) sequencing to analyze circulating T cells from a multi-center cohort of ICI-M patients pre/post therapy, in comparison to irAE and non-irAE cancer controls (Fig. S1). Combining with spatial transcriptomics of the affected muscle, we identified irAE specific CD38^hi^ CD8 T cells expanded in blood and muscle of ICI-M patients. Moreover, abatacept altered the composition and clonality of CD8 T cells, pinpointing a dynamic link between CD38^hi^ CD8 T cell and clonal KIR^+^ CD8 Treg subset. These results provide evidence for monitoring specific immune activation and regulation at critical clinical decisions points.

## Results

A multi-center cohort of 26 ICI-M patients were analyzed, including 25 patients receiving PD-1/PD-L1 and 1 receiving PD-1/CTLA-4 combinational therapy. 24 patients presented with grade 3~4 ICI-M. All received glucocorticoids, averaging maximus 285 mg/day of methylprednisolone equivalent. While receiving similar steroids dosage, 15 out of 16 patients treated with abatacept survived (93.75%), compared to 5 of 10 without abatacept (50%). Detailed clinical information was summarized in Table S1.

For unbiased discovery, scRNA-seq of peripheral blood of ICI-M (n=2) and ICI-A (n=3) patients were performed. 12 distinct clusters comprising of 12340 T cells were observed (Fig. 1a) with distinct transcriptomic features (Fig. S2a and Table. S2). CD8 T subsets C1, C3, C4, C6 were enriched in ICI-M while CD4+T subsets C0, C2, C8, C9 expanded in ICI-arthritis (Fig. 1b). Four cytotoxic CD8 subsets enriched in ICI-M showed distinct transcriptomic patterns (Fig. 1c and S2b). C3 and C6 exhibited elevated levels of GZMK, HLA-DRA and CD38. C4 expressed GZMB, GZMH, KIR3DL1, and KIR2DL1. C1 expressed high level of IL7R and moderate level of GZMB and GZMH. Differentially expressed genes (DEGs) in C3 and C6 were enriched in T cell signaling pathway, cell adhesion molecules and viral myocarditis related pathway which were down regulated in C1, while C4 showed enrichment in cytotoxicity, antigen processing and presentation (Table. S3). TCR analysis showed significantly higher clonality in C1, C3, C4, and C6 in ICI-M (Fig. 1d and S4), and high similarity was observed between C3 and C6 (Fig. S2c). Based on a set of signature genes we previously identified in ICI-A^12^, C3 and C6 were the subsets most closely resembling the irAE-specific CD38hi population directly bound and activated by ICI (Fig. S2c). Hyperexpanded and large clonal TCR sequences were grouped using GLIPH2 into 35 clusters, of which 18 specific to ICI-M and 17 to ICI-A—indicating distinct antigen recognition (Table. S4). While single clones recognizing specific viruses and human antigens (p53, MLANA in ICI-A donors) were found, large and expanded clones were not matched to reported autoantigen in VDJdb (Table. S5).

**Figure 1.**
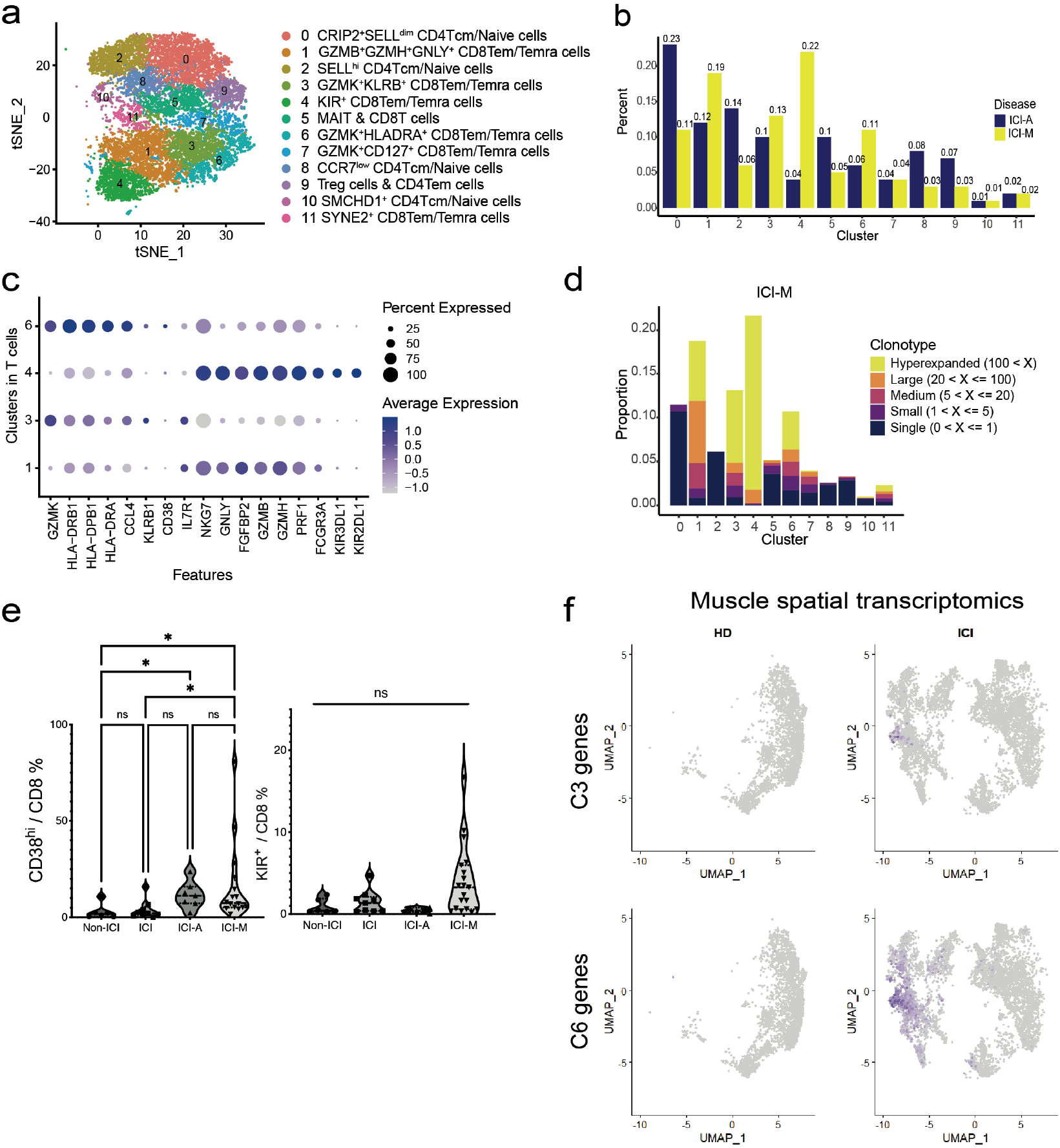
Extensive clonal expansion of CD38^hi^ and KIR^+^ cytotoxic CD8 T cells in ICI-myotoxicity. a). tSNE visualization of circulating T cells from ICI-M (n=2) and ICI-A (n=3) forming 12 distinct clusters. b). Frequency of T cells clusters. c). Dot plot of DEGs that distinguish C1, C3, C4 and C6. Dot color represents the average expression. Dot size represents the percentage of expressing cells. d). The proportion of cells in each cluster from ICI-M. Color indicates TCR clonotype. e). Frequency of CD38^hi^ cells and KIR^+^ cell among CD8 T cells from ICI-M (n=16), ICI-A(n=7), ICI-treated without irAE (ICI, n=9), and non-ICI controls (non-ICI, n=6). f). UMAP visualization of C3/C6 signature genes using spatial RNA-seq of muscle tissue from healthy donor (n=5) and ICI-M (n=5).

To identify irAE-specific transcriptomic changes, integration of public non-irAE control data^11^ confirmed that C3/6 and C4 were represented only in ICI-M and ICI-A, while C1 was enriched in controls (Fig. S3a-b). Using previously validated protein markers and gating strategy^12^ (Fig. S5), multicolor flow cytometry validated significant expansion of CD38hi CD8 T cells in ICI-M over non-ICI (8-fold) and ICI-non-irAE (5-fold) controls (Fig. 1e). Using C3/6 signature genes, these populations were clearly visualized in ICI-M affected muscle and not found in healthy donors (Fig. 1f). KIR^+^ CD8 T cells showed increased proportion in ICI-M blood (2-4-fold over controls) but were scarce in muscle tissue.

Using established protocols^6,7^, tailored abatacept dosing based on CD80/86 blockade, in addition to corticosteroids +/- ruxolitinib successfully rescued clinically fulminant ICI-M in a multi-center cohort with 93.75% three-month survival. (Fig. 2a, Table S1). T cells from paired blood before and after abatacept showed altered composition and clonality of CD8 T cells in a week (Fig 2b-c). C4 KIR+ CD8 T cells had the highest proportion of top clonotypes shared before and after abatacept, which was further expanded as the patient recovered clinically. A lower level of clonal expansion was observed in C1, C3 and C6, while C3 CD38-like CD8 T cells showed a decreased proportion in top shared clonotypes (Fig. 2c). As ICI-M patients (n=12) responded to abatacept clinically comparing to baseline, the frequency of CD38^hi^ C3/6 decreased to an average of 0.5-fold, while KIR+ C4 expanded to an average of 1.7-fold (Fig. 2d, Fig. S6a-d). CD38^hi^ CD8 T cells did not decrease in patients treated with steroids alone (n=4). KIR^+^ CD8 T cells frequency did not increase in steroids alone treated patients (n=3), and it did not increase upon follow-up at 3 weeks to 2 months after patients discontinued abatacept, suggesting a transient treatment response specific to abatacept (Fig. S6e). Cell-cell communication analysis among T cells revealed C4 KIR+ CD8 T cells have robust connections with C1, C3 and C6 (Fig. 2e). In addition to HLAs-CD8 and HLAs-KLRs ligand-receptor pairs, KIRs on C4 make contributes to communication with C3 and C6 (Fig. 2f). DEGs between C4 and C3/6 highlighted KIR molecule enrichment in C4 and tissue-inflaming GZMK in C3/6 (Fig. 2g). Pathway analysis revealed enrichment of genes involved in T cell activation, cell adhesion and MHC mediated antigen presentation in C3/6, and cell killing in C4 (Fig. 2h), suggesting C4 may serve as a regulatory population suppressing inflammatory C3 and C6 clusters.

**Figure 2.**
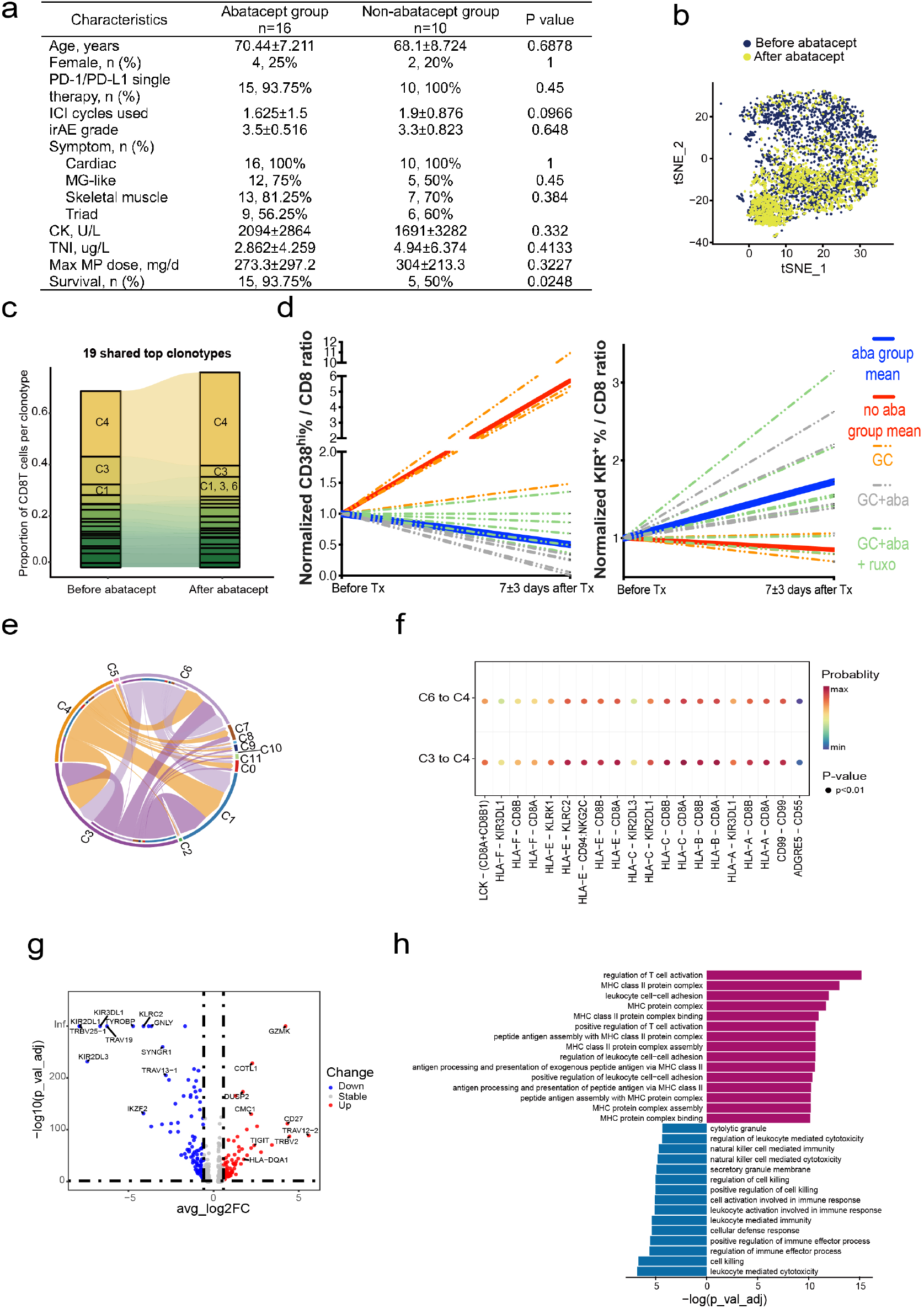
Abatacept altered the frequency and clonality of potentially interacting CD38^hi^ and KIR^+^ cytotoxic CD8 T cells in ICI-myotoxicity. a). Clinical characteristics of ICI-M patients. b). tSNE visualization of circulating T cells before and after abatacept. c). The proportion of CD8 T cells clusters before and after abatacept. d). Normalized frequency of CD38^hi^ cells (n=16) and KIR^+^ cell (n=15) among CD8 T cells from ICI-M before and after treatment. The baseline frequency before treatment was normalized to 1 and the frequency from the same donor after treatment was normalized as the ratio of baseline. Solid blue line represents the average value of patients receiving abatacept. Solid red line represents the average value of patients receiving glucocorticoids. Dotted grey line represents patients (n=5) treated with glucocorticoids and abatacept. Dotted green line represents patients (n=7) treated with glucocorticoids, abatacept and ruxolitinib. Dotted orange line represents patients (n=3-4) treated with glucocorticoids. GC: glucocorticoids. Aba: abatacept. Ruxo: ruxolitinib. e). Chord plot of the inferred cell-cell communication network among T cell clusters. Color indicates cluster identity. Chord width represents the interaction strength. f). Dot plot of significant (p<0.01) ligand-receptor interaction pairs between C3-C4 and C6-C4. Circle color represents communication probability. g). Volcano plot of DEGs between C3/6 (red) and C4 (blue). h). Pathway enrichment analysis of DEGs between C3/6 (red) and C4 (blue).

In summary, scRNA-seq and scTCR-seq revealed prominent clonal expansion of CD38^hi^ CD8 T cells and KIR^+^CD8 T regulatory population. Flow cytometry verified increased CD38^hi^ CD8 T cells in the blood and muscle tissue of fulminant ICI-M compared to controls. As patients improve clinically to abatacept, two potentially interacting populations, CD38^hi^ CD8 inflammatory T cell frequency decreased while KIR^+^CD8 regulatory T cell population clonally expanded. Dynamic monitoring of these populations could assist rapid disease recognition, real-time therapeutic assessment, and provide insights into irAE pathogenesis.

## Discussion

In this study of multi-center cohort, we observed clonal expansion of two functionally distinct cytotoxic CD8 T cells in ICI-myotoxicity. Patients who received abatacept demonstrated improved survival of this life-threatening irAE, coinciding with a decrease in CD38^hi^ CD8 T cells and a clonal increase in KIR^+^CD8 T cells. Dynamically analyzing these two subsets as biomarkers offers a unique opportunity to evaluate therapeutic response and provides novel insights into the mechanism of immune dysregulation in immunotherapy and autoimmunity.

The ICI-M patients in this study demonstrated early onset of severe cardiac, respiratory and skeletal muscle involvement, attesting the importance of early awareness and comprehensive assessment for patients receiving ICI for the first few cycles. Despite similar severity and steroid dosage, patients receiving a precision-dose of abatacept demonstrated improved survival supported by real-time analysis of T cell dynamics and CD80/CD86 blockade. We did not observe additional T cell trend in patients receiving combinational abatacept and ruxolitinib, more analysis is needed in more patients to investigate whether ruxolitinib provide additional therapeutic benefits.

Building upon our prior identification of CD38hi cytotoxic CD8 T cells bound by anti-PD-1 antibody in ICI-arthritis^12^, this study establishes these cells as critical indicators of immune activation and tissue inflammation in ICI-myotoxicity (ICI-M). Their significant expansion in ICI-M patients compared to non-irAE and non-ICI controls enables physicians to dynamically gauge disease severity and monitor real-time responses to immunosuppressive therapy. The pathogenic role of these cells is supported by recent findings of CD38’s pro-survival function in inflamed tissues^15^ and their enriched expression of granzyme K, a shared tissue-inflammatory program in autoimmunity and cancer^16^. Furthermore, these CD38hi cells are enriched for CXCR3, suggesting a mechanism for myocardial infiltration via interaction with CXCL9/10+ macrophages, a pathway previously shown to exacerbate cardiac injury^81117^. The CD38^hi^ cytotoxic CD8 T in this study are enriched for CXCR3 (Fig. S5). Similar mechanism could be responsible for these cells’ migration towards the muscle tissue to exert inflammatory effects.

Additionally, our study identified clonal expansion of KIR^+^ regulatory CD8 T cells in ICI-myotoxicity, with their numbers transiently increasing in tandem with a positive therapeutic response. Previous studies have shown the correlations between KIR+CD8 T cells and cancer immunosurveillance^14,18^ and autoimmune diseases such as systemic lupus erythematosus^13^, while our study is the first to identify this population in irAEs. Prior research have demonstrated that KIR^+^CD8 T cells exhibit a restricted TCR repertoire, aligning with the results of our investigation. Similar to murine Ly49+CD8+ T cells, which suppress self-reactive CD4+T cells, KIR^+^CD8 T cells were studied in autoimmune disease to directly kill self-reactive pathogenic CD4^+^T cell via cytotoxic molecules and to induce apoptosis of pathogenic CD4^+^T in a contact-dependent manner, with the involvement of MHC-I molecules in both processes^13^. Supporting an antigen-specific, context-dependent regulatory role, tumor-antigen-specific KIR^+^ CD8 T cells shared between tumor and blood were found to target tumor-peptide reactive CD8^+^ T cells, correlating with poor overall survival in melanoma^18^ and liver cancer^14^. KIR+ CD8 T cells we found in ICI-myotoxicity also highly express cytotoxic molecules GZMB and PRF1, while cell-cell communication analyses suggest they interact with potential pathogenic T cell clusters through MHC-KIR pairs, suggesting they may serve as a negative feedback mechanism to mitigate pathogenesis by eliminating autoreactive T cells through cytotoxic activity. Further experiments in vivo and in vitro in an antigen-specific setting, as well as low-grade ICI-myotoxicity patient who may have experienced this self-regulating process, is needed to confirm this hypothesis.

In conclusion, the frequency and clonality of CD38^hi^ cytotoxic CD8 T cells and KIR^+^ regulatory CD8 T cells indicate disease severity and therapeutic response in ICI-myocarditis. Analyzing these populations enhances our understanding of ICI-M pathogenesis and offers a potential strategy for personalized treatment of life-threatening irAEs.

## Data Availability

All data produced in the present study are available upon reasonable request to the authors

## Acknowledgements

This work has been supported in part by funding from National Natural Science Foundation of China Grant (82402095 to RW), Science and Technology Innovation Plan of Shanghai Science and Technology Commission (23YF1423000 to RW).

---

**Supplementary figure 1.**
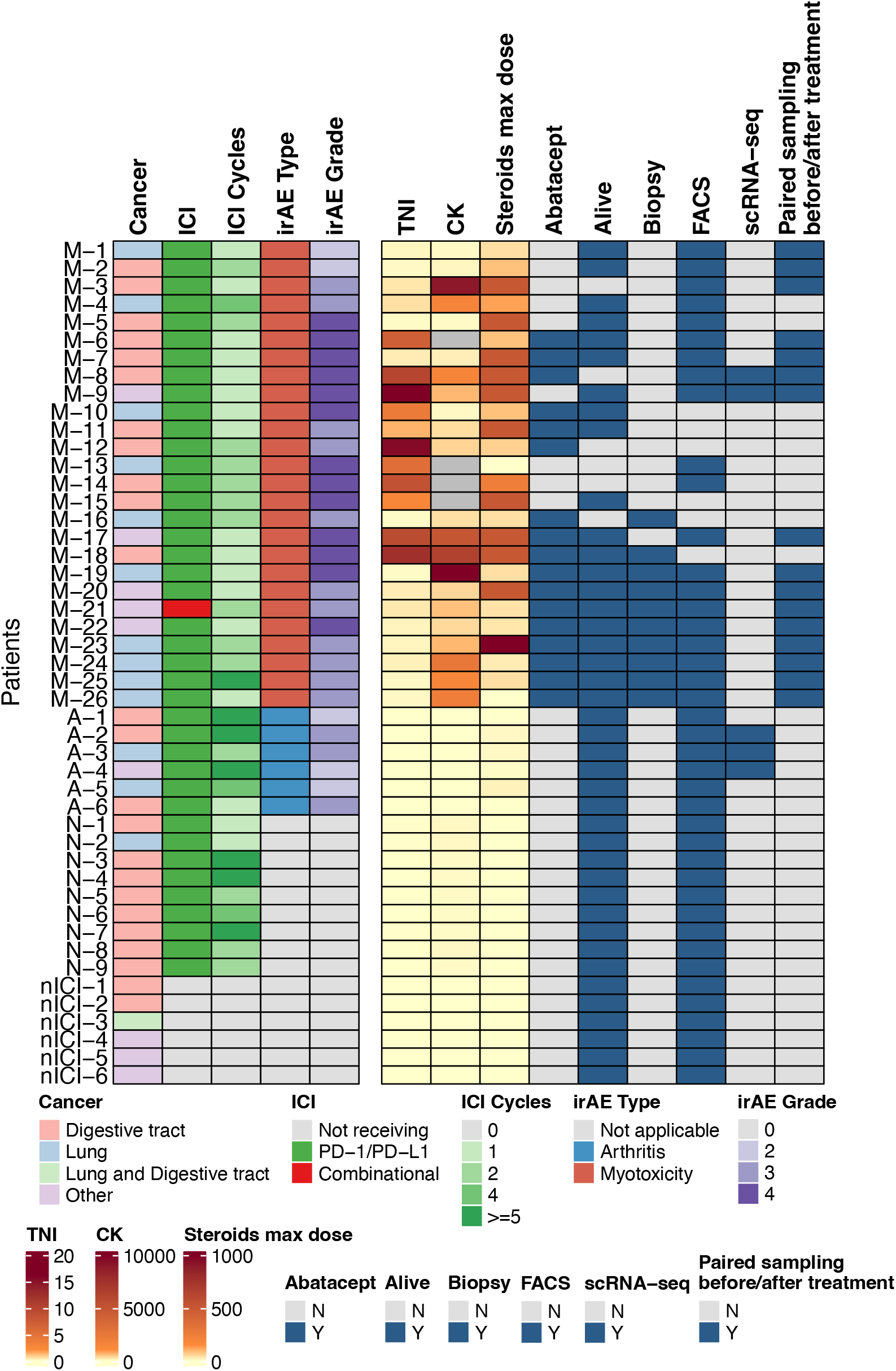
Summary of patient characteristics. M1-26, ICI-myotoxicity donors. A1-6, ICI-arthritis donors. N1-9, cancer patients receiving ICI without developing irAE. nICI1-6, cancer patients without receiving ICI. TNI, serum troponin level. CK, serum creatine kinase level. Steroids max dose, mg/d in methylprednisolone equivalent. Alive, survival status in three months.

**Supplementary figure 2.**
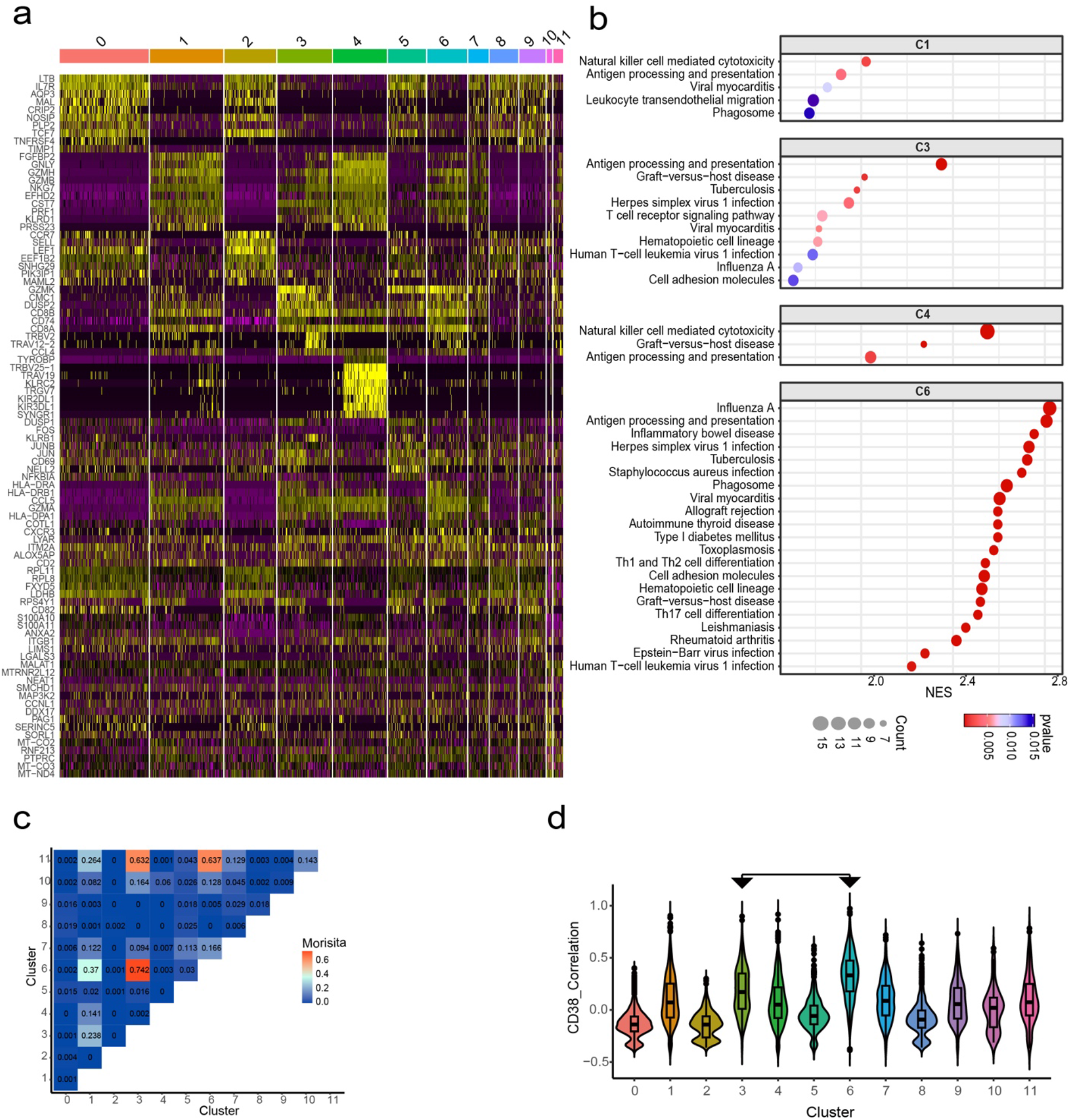
Transcriptomic features of T cell clusters. a). Heatmap of differentially expressed genes in each T cell clusters. b). Pathway enrichment analysis of differentially expressed genes in C1, C3, C4 and C6. Dot size represents gene count. Color represents p value. c). Plot representing the Morisita index value for each comparison showing the repertoire overlap. Color represents Morisita index value. d). Correlation of CD38 signature score in each cluster.

**Supplementary figure 3.**
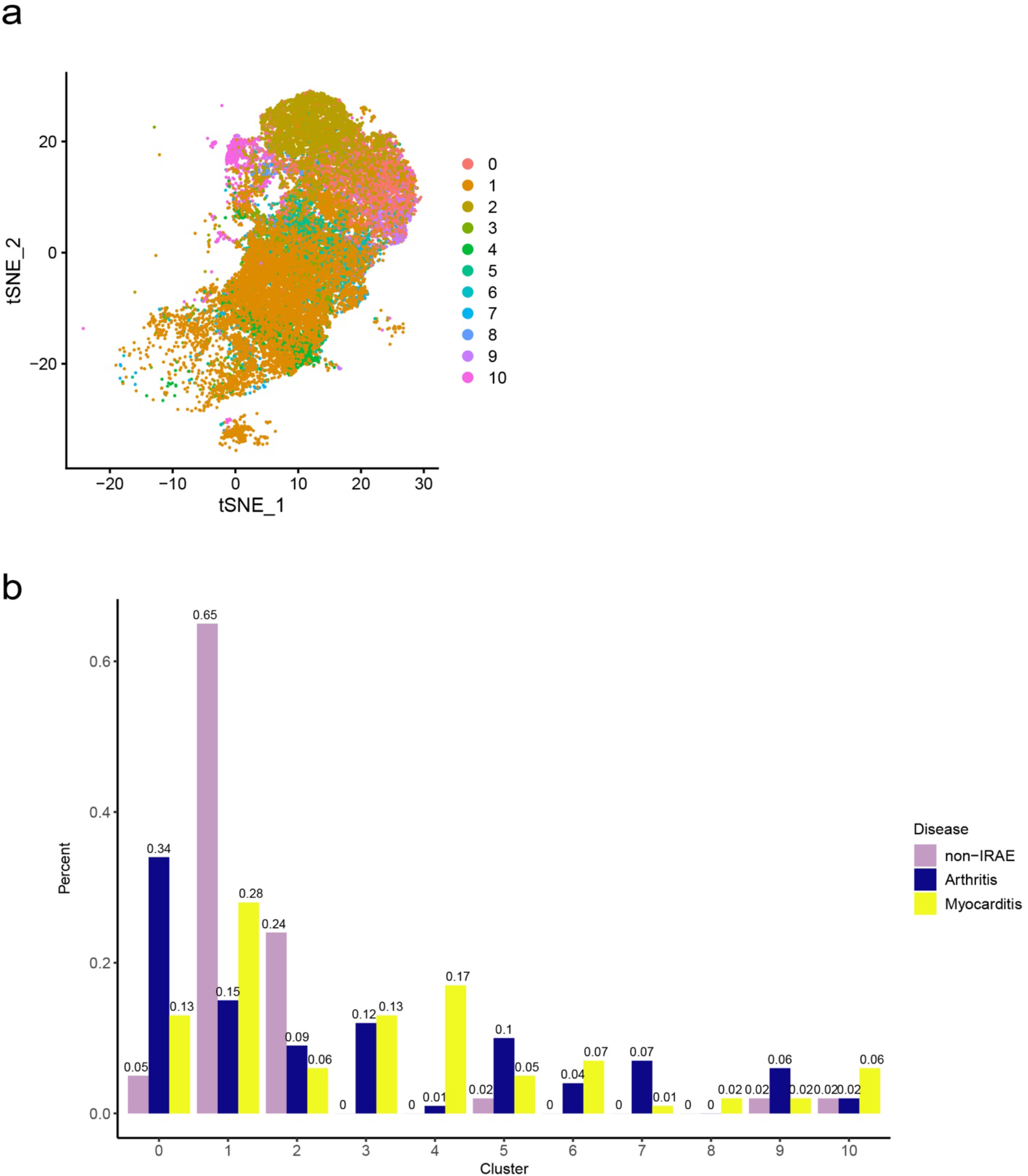
Integration of public scRNA-seq data reveals irAE specific T cell clusters. a). tSNE visualization of circulating T cells from public data of non-irAE patients treated with ICI (n=3), and scRNA-seq data in figure 1. b). Frequency of clusters of non-irAE, ICI-M and ICI-A T cells.

**Supplementary figure 4.**
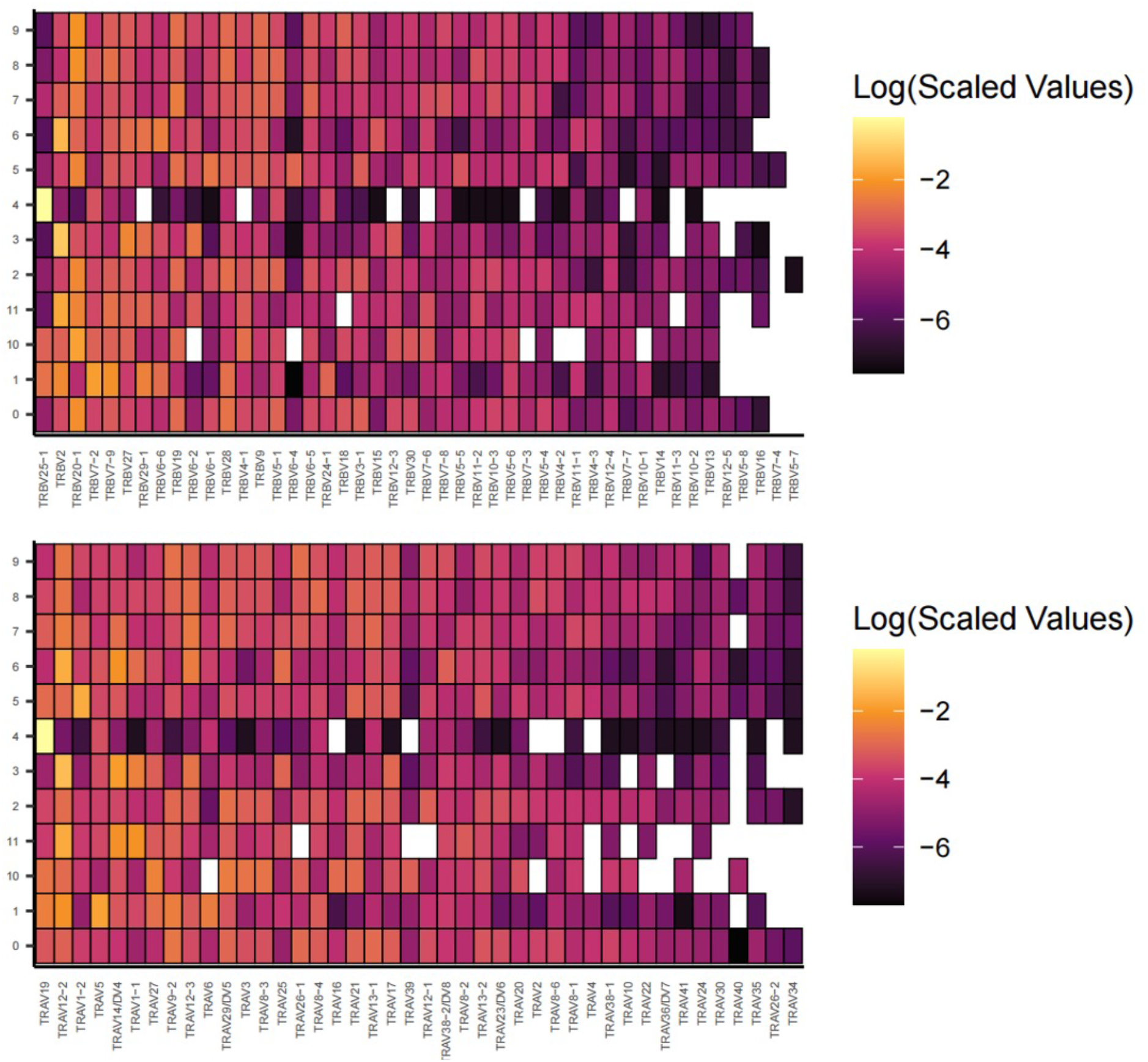
TRBV and TRAV usage in T cell clusters.

**Supplementary figure 5.**
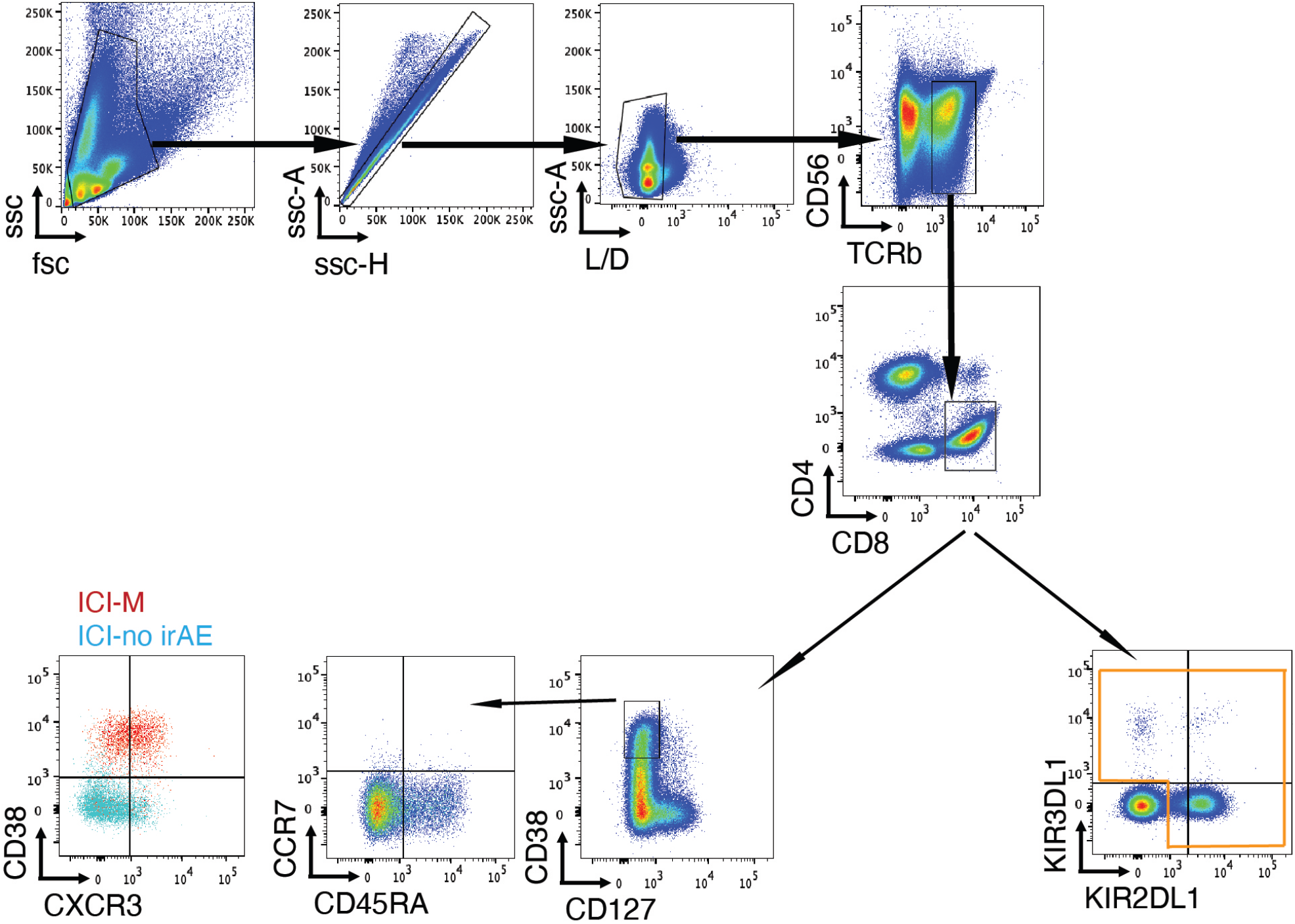
Gating strategy for CD38^hi^ CD8 T cells and KIR^+^ CD8 T cells. CD38^hi^ CD8 T cells were mostly CCR7^-^ effector memory cells, and these cells from ICI-M donors were enriched for CXCR3 over ICI-no irAE controls. KIR^+^ CD8 T cells were counted as the combined KIR3DL1^+^ and KIR2DL1^+^ cells.

**Supplementary figure 6.**
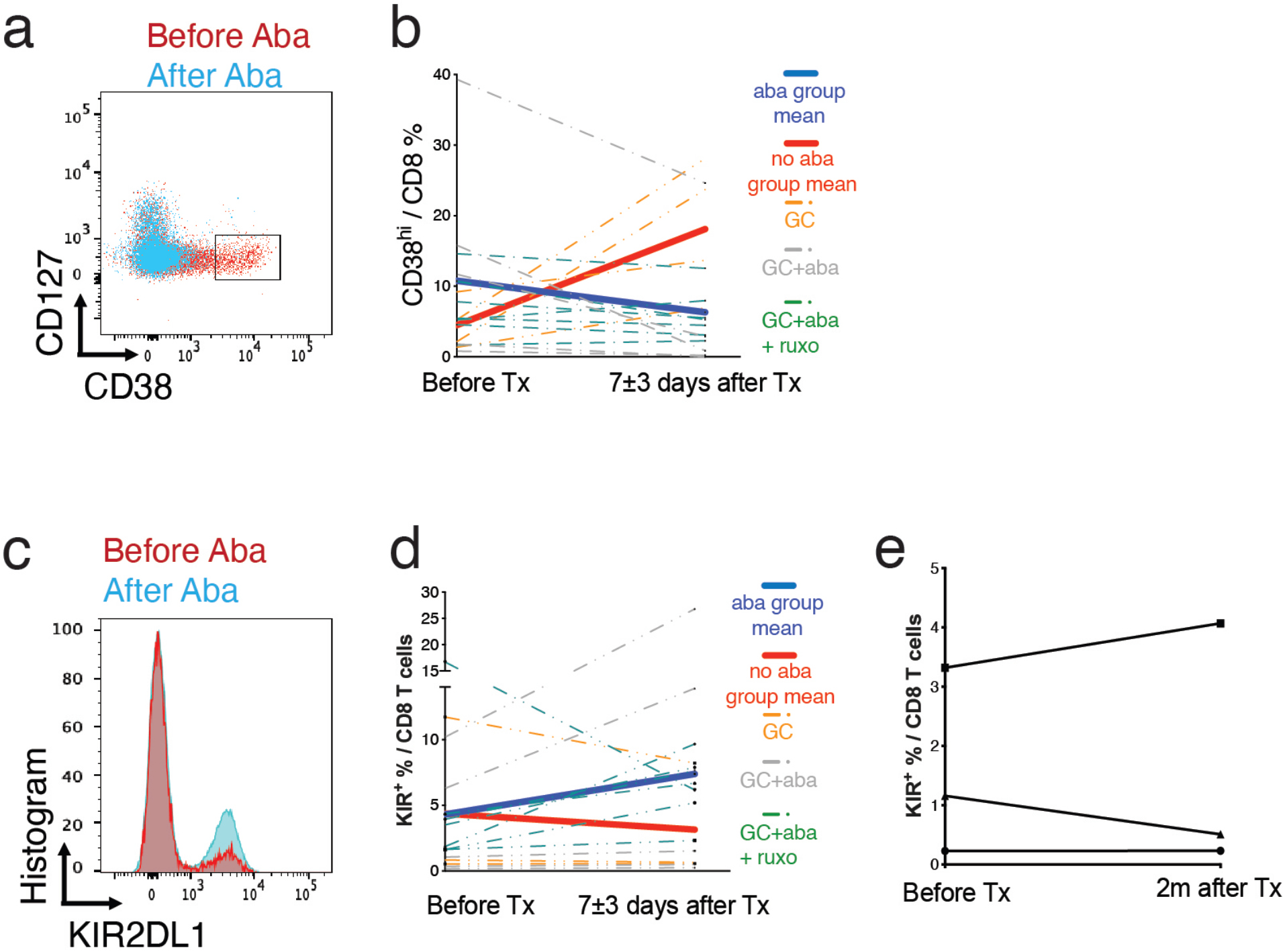
Dynamic changes of CD8 T cell subsets after treatment in ICI-M patients. a.) Representative flow cytometry plot of CD38^hi^ CD8 T cells before and after abatacept treatment. b). Frequency of KIR^+^ cell among CD8 T cells from ICI-M (n=16) before and after treatment. c.) Representative flow cytometry plot of KIR^+^ CD8 T cells before and after abatacept treatment. d). Frequency of KIR^+^ cell among CD8 T cells from ICI-M (n=15) before and after treatment. e). Frequency of KIR^+^ cell among CD8 T cells from patients (n=3) before and after 2 months of abatacept. In b) and d), solid blue line represents the average value of patients receiving abatacept. Solid red line represents the average value of patients receiving glucocorticoids. Dotted grey line represents patients (n=5) treated with glucocorticoids and abatacept. Dotted green line represents patients (n=7) treated with glucocorticoids, abatacept and ruxolitinib. Dotted orange line represents patients (n=3 ~4) treated with glucocorticoids. GC: glucocorticoids. Aba: abatacept. Ruxo: ruxolitinib.

## Online Methods

### Study approval and patient samples

Biological samples were collected with informed consent. Retrospective study and sample collection was approved by Renji Hospital Shanghai Jiao Tong University (China) and Pitié-Salpêtrière, APHP–Sorbonne University (Paris, France). irAEs were diagnosed by experienced specialists according to established standards and protocol^19,20^. Peripheral blood samples were collected from patients in a multi-center cohort of ICI-M (n=26, PBMC available in n=20), ICI-A (n=7), ICI-without irAE (ICI, n=9), and cancer-without ICI therapy (non-ICI, n=6) controls (supplementary table 1). ICI-M patients were managed with standard of care, and the choice of immunosuppressants was determined by the medical team in charge. The dose of abatacept was titrated based on CD80/CD86 blockade for ICI-myotoxicity according to established protocol^6^. serial paired blood samples before and after abatacept were collected for analysis. Muscle samples from ICI-M patients (n=5) and healthy donors (n=5) were collected for transcriptomic analysis.

### Peripheral blood sample preparation

PBMC from patients were obtained by density centrifugation using Ficoll-Paque Plus (Cytiva, USA) and preserved in liquid nitrogen for batched analyses. Cryopreserved cells were thawed into RPMI medium + 10% fetal bovine serum when experimental analyses. Dead cells in single-cell suspension were removed using Dead Cell Removal Kit (Miltenyi Biotec, Germany). After that, T cells were isolated by magnetic-activated cell sorting using CD3 microbeads (Miltenyi Biotec, Germany) following the instructions and then passed through 35 µm nylon mesh for downstream sequencing.

### Muscle sample spatial RNAseq processing

Muscle tissue was collected for diagnosis purpose and frozen in pre-cooled isopentane and stored at −80°C. Sections from muscle samples for ten patients (n=5 for ICI-M and 5 for healthy donors) were processed following the manufacturer’s recommendation using Visium Spatial Gene Expression kit (10x Genomics, Pleasanton, CA, USA). Visium libraries were sequenced using a NextSeq Illumina sequencer at a sequencing depth going from 40M to 100M read-pairs per sample depending on the section size. Alignment and mapping were done using SpaceRanger v1.0.0 (10X Genomics) and used the same packages as single cell transcriptome analysis.

### Single cell sequence library preparation

5’ gene expression and paired TCR libraries were prepared using 5’ Chromium Next GEM Single Cell v2 (Dual Index) following the kit instructions. The pooled 5’ gene expression and TCR libraries were sequenced using Illumina NovaSeq6000 instrument by Genergy Bio-technology Co. Ltd (Shanghai) according to 10X Genomics guideline.

### Single cell transcriptome analysis

Cell Ranger was used to process FASTQ files and align the reads to human reference genome GRCh38. Seurat package (v5.0.2) was used to perform downstream analysis. Quality control (QC) was performed to remove poor quality cells that had less than 200 genes, more than 2500 genes, or > 5% mitochondrial gene expression. Genes expressed in less than 3 cells were excluded. We got a total of 32597 cells and 21568 genes for further analysis. Data was log-normalized and scaled with default parameters. Principle component analysis (PCA) was used with 2000 highly variable genes. Batch effect was corrected by Harmony (v1.2.0) to improve integration and cell cycle score was performed. 29 clusters were found at 1.9 resolution. Cell types were auto-annotated using CellTypist package (v2.0) with Adult_Human_Blood and Immune_All_Low reference, and then manual verified based on differentially expressed genes (DEGs) (Table. S2) found by FindAllMarkers function. T cell populations were extracted for QC again (QC standards seeing in TCR sequencing analysis) and then go further analysis. The AddModuleScore function was used to calculate CD38 correlation scores, CD38 correlated gene set (Table. S3) was from Wang et al., Science Immunology 2023^12^.

### Cell-cell communication analysis

Cell-cell communication prediction of potential receptor-ligand pairs among T cells was performed by cellchat. The human cellchat database was used as reference, which considers the known composition of ligand-receptor complexes, including complexes with multimeric ligands and receptors, as well as cofactors like soluble agonists, antagonists, co-stimulatory and co-inhibitory membrane-bound receptors. Potential bias caused by population size was removed. For calculation the communication strength (or probability), each cell type was labeled and randomly arranged 100 times. Statistically significant communications were identified as p value<0.05.

### TCR sequencing analysis

Cell Ranger vdj pipline was employed to generate clonotype data from VDJ FASTQ files. These reads were mapped to GRCh38 human reference sequence. scRepertoire (v2.0) package was used for integrating TCR sequencing with RNA sequencing. QC was performed as following, 1) isolating the top 2 expressed chains in cell barcodes with multiple chains; 2) retaining cells that both with TCR information and RNA information. We got a total of 13445 T cells, and 7544 unique clonotypes, where a unique clonotype share the same CDR3 alpha and beta sequences. Clonotype details were incorporated into the metadata of Seurat object by combineExpression function, facilitating the investigation of the relationship between TCR information and its phenotype. Morisita index, an ecological measure of the dispersion of individuals within a population, incorporating the size of the population, was used to measure of similarity of clusters. GLIPH2, an algorithm to cluster TCR sequences that could recognize the same antigen, was used to group TCRs with reference version 2.

### Flow cytometry staining

A 9-color panel of antibodies includes anti-TCRb-FITC (IP26), anti-CD4-BUV395 (RPA-T4), anti-CD8-BUV496 (RPA-T8), anti-CD38-PerCP/Cy5.5 (HIT2), anti-CD127-APC (A019D5), anti-KIR3DL1-BV421 (DX9), anti-KIR2DL1-PE (HP-DM1), anti-CXCR3-AlexaFluor700 (G025H7), all from BioLegend and LIVE/DEAD Fixable Aqua Dead Cell Stain Kit from Invitrogen. Cryopreserved cells were thawed and counted. Cells were stained in antibody cocktails at 4 °C for 20 minutes and washed twice with FACS buffer. Then cells were passed through a 70 μm filter and detected fluorescence intensity using BD Fortessa. Data was analyzed by FlowJo 10.8.1.

### Statistics

Analyses were performed using R 4.2.0 and GraphPad Prism 8.0 software. Multigroup comparison of flow cytometry data was performed by Kruskal-Wallis followed by Dunn’s multiple comparison test for subgroup comparison. P value < 0.05 was considered statistically significant after adjusting. Other significant differences were determined by the algorithm default of R packages.

## Notes

### Competing Interest Statement

The authors have declared no competing interest.

### Author Declarations

Study and sample collection were approved by Ethics Committee of Renji Hospital, Shanghai Jiao Tong University (identification no. 20242 35 C)

